# Effects of Non-Surgical Periodontal Therapy on Dental Plaque Microbiome

**DOI:** 10.64898/2026.06.30.26356898

**Authors:** Qingguo Wang, Bing-Yan Wang, Derek Wilus, Hua Xie

## Abstract

Periodontitis, a chronic inflammatory disease affecting approximately 40% of U.S. adults aged 30 years and older, is characterized by dysbiosis of the dental plaque microbiome. However, although scaling and root planing (SRP) is the cornerstone of periodontal treatment, its effects on the taxonomic composition and functional potential of the dental plaque microbiome remain incompletely understood. In this study, we used whole-metagenome shotgun sequencing to characterize taxonomic composition and functional potential in dental plaque microbiomes collected from 39 patients with Stage II or III generalized periodontitis before and 3-4 months after SRP. Consistent with clinical improvement, periodontal therapy significantly reduced bleeding on probing and plaque index. Whole-metagenome shotgun sequencing identified 3.18 million non-redundant genes and 12,353 microbial species across 78 samples, revealing increased gene and species richness after treatment, along with a significant restructuring of microbial community. Established periodontal pathogens, including *Porphyromonas gingivalis* and *Tannerella forsythia*, as well as the emerging pathogen *Escherichia coli*, decreased following treatment, whereas health-associated early colonizers, including multiple *Actinomyces* species and *Streptococcus cristatus*, increased. Functional annotation using the Carbohydrate-Active Enzymes (CAZy) database identified treatment-associated differences in several carbohydrate-active enzymes, including multiple glycosyltransferases, indicating remodeling of the predicted functional potential of the dental plaque microbiome. These findings demonstrate that successful SRP promotes coordinated taxonomic and predicted functional remodeling of the dental plaque microbiome and highlight the value of shotgun metagenomic sequencing for characterizing both taxonomic and functional recovery following periodontal therapy.

## 1. INTRODUCTION

Periodontitis is the 6^th^ most common non-communicable disease worldwide and a major cause of tooth loss in adults [1]. Despite substantial advances in prevention and treatment over the past five decades, periodontitis continues to affect a large proportion of the global population [2, 3]. According to the National Health and Nutrition Examination Survey 2009-2014, approximately 40% of adults aged 30 or older in the US had one stage of periodontitis [4]. The disease is initiated and sustained by a dysbiotic dental plaque biofilm that triggers a chronic host inflammatory response and may lead to progressive destruction of the teeth’s supporting structures.

Periodontal therapy includes scaling and root planing (SRP), which disrupts and removes dental plaque to reduce periodontal inflammation. These non-surgical periodontal treatments are the cornerstone of periodontal treatment and are often effective in reducing periodontal pocket depth (PD) and improving clinical attachment levels (CAL). For patients with more advanced disease, SRP may be supplemented with adjunctive therapies, including systemic or local antimicrobials and periodontal surgery. Numerous studies have demonstrated that SRP, either alone or in combination with adjunctive therapies, suppresses periodontal pathogens and promotes clinical improvement [5–8].

Previous investigations using checkerboard DNA-DNA hybridization and quantitative PCR demonstrated significant reductions in periodontal pathogens following periodontal therapy [5–8]. Importantly, microbial responses were not homogeneous, with some taxa showing rapid reductions after treatment, whereas others exhibited delayed responses or little change, suggesting complex ecological restructuring of the oral microbiome following therapy [5]. Recently, 16S rRNA gene sequencing studies have reported substantial shifts in microbial community composition following treatment, with post-treatment microbiomes resembling those observed in periodontally healthy patients [9, 10]. However, recent evidence indicates that microbiome dysbiosis may persist even after successful clinical remission of periodontal disease, suggesting that conventional treatment may not fully restore the oral microbial ecosystem [7]. In addition, emerging evidence has implicated several previously underappreciated species, including *Filifactor alocis*, *Enterococcus faecalis*, *Haemophilus influenzae*, and *Escherichia coli*, as potential contributors to periodontal disease [8].

Despite these advances, most previous studies relied on targeted molecular approaches or 16S rRNA sequencing, which provide limited taxonomic resolution and little information regarding microbial functional capacity. Consequently, the effects of periodontal therapy on species-level microbial composition and predicted functional potential remain incompletely understood.

Whole-metagenome shotgun sequencing enables species-level taxonomic profiling while simultaneously providing insight into the predicted functional potential of microbial communities [11, 12]. In this study, we applied whole-metagenome shotgun sequencing to characterize the taxonomic composition and predicted functional potential of the dental plaque microbiome before and after non-surgical SRP in patients with periodontitis. Our objective was to identify treatment-associated taxonomic and functional changes and to provide a more comprehensive understanding of dental plaque microbiome remodeling following periodontal therapy.

## 2. MATERIALS AND METHODS

### Study cohorts

The research protocol was approved by the Committee for the Protection of Human Subjects at the University of Texas Health Science Center, Houston (IRB number: HSC-DB-17-0636) [11, 12].

Participants were recruited during routine dental visits in a clinic in the School of Dentistry, University of Texas Health Science Center, Houston, between 2017 and 2022. Clinical oral examinations were conducted by trained dental examiners, who were also faculty members of the School of Dentistry. The examiners are calibrated annually in the diagnosis of periodontitis. Individuals aged 21-75 years were screened at baseline (before any periodontal treatment) after the initial periodontal examination, including determination of the plaque index (PI) and bleeding on probing (BOP) [13]. Radiographs were conducted to assess bone loss. All enrolled participants were diagnosed with Stage II or III generalized periodontitis, regardless of their grade, based on the 2017 World Workshop Classification [14, 15]. The enrolled patients also met the following criteria: ≤4 tooth loss due to periodontitis, interdental CAL ≥3 mm and PD ≥5 mm at two or more teeth in different quadrants, and radiographic bone loss ≥15%. Other criteria for study participation were as follows: 1) no SRP within the previous year or periodontal surgeries in the previous five years; 2) no antibiotic therapy in the previous six months; 3) no pregnancy. Information on demographics and self-reported dental and medical histories of the participants were obtained from the electronic health records.

### Dental plaque sample collection

Dental plaque samples, including supra and subgingival dental plaques, were collected at baseline and three or four months after non-surgical SRP treatment by board-certified periodontists using sterile paper points prior to any dental treatment and labeled numerically according to the sampling sequences. The paper points were placed in ≥5mm pockets in different quadrants for 1 minute, then immediately immersed in an Eppendorf tube containing 0.5 ml of Tris-EDTA (TE) buffer (pH 7.5) [16]. Bacterial pellets were harvested by centrifugation and then resuspended in 100 µl TE buffer. Samples were stored at −80 °C until use.

### Library construction, quality control, and sequencing

To perform shotgun metagenomic sequencing, dental plaque samples were shipped on dry ice to Novogene Co., Ltd. (Sacramento, CA, USA). Genomic DNA from the samples was randomly sheared to an average fragment size of approximately 350 bp using a Covaris ultrasonic disruptor. Sequencing libraries were subsequently prepared via end repair, A-tailing, adapter ligation, purification, and PCR amplification. Fragment integrity and insert size distribution of the resulting libraries were evaluated using an Agilent/AATI Fragment Analyzer. Libraries meeting the target insert size criteria were quantified using qPCR; only those with an effective concentration >3nM were advanced for sequencing.

A total of 78 quality-validated metagenomic libraries were pooled and sequenced on the Illumina NovaSeq platform using a 150 bp paired-end (PE150) strategy. High sequencing quality and base-calling accuracy were maintained across all runs, with Q20 and Q30 quality scores consistently exceeding 96% and 91%, respectively.

### Data preprocessing

The raw sequencing reads were preprocessed using the fastp software (version 0.23.1) to remove low-quality sequences and adapters [17]. Paired-end reads were discarded if either read met any of the following criteria: (i) contained adapter contamination; (ii) contained more than 10% ambiguous nucleotides (N); or (iii) contained more than 50% low-quality bases with a Phred quality score below 5. To eliminate host DNA contamination, clean reads were aligned to the human reference genome, and all reads that mapped to it were filtered out. Bowtie2 (version 2.2.4) was used for host read filtering with the parameters --end-to-end, --sensitive, -I 200, and -X 400 [18]. The resulting non-host reads were retained for subsequent metagenomic analyses.

### Gene prediction and abundance analysis

Clean reads were assembled into contigs using MEGAHIT (version 1.2.9)[19]. Contigs shorter than 500 bp were removed before gene prediction. The software MetaGeneMark was used in default settings to predict open reading frames (ORFs) from assembled contigs [20]. ORFs shorter than 100 nucleotides were excluded. Then, these ORFs underwent dereplication using CD-HIT (version 4.5.8) to create non-redundant gene catalogues [21, 22]. Clean data were then mapped to the gene catalogues using Bowtie2 to quantify gene abundance. This abundance was calculated based on the number of mapped reads and gene length, using the following formula:

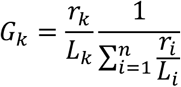

where *r* represents the number of mapped reads and *L* represents gene length. Downstream analyses were performed based on the abundance of the gene catalogues.

### Species annotation

Taxonomic annotation was performed using DIAMOND software (version 0.9.9.110) [23] to align predicted protein sequences against the MicroNR database (version 2024.03), which contains bacterial, archaeal, fungal, and viral sequences derived from the NCBI non-redundant (NR) database. Sequence alignments were conducted using the blastp algorithm with an E-value cutoff of *1 × 10^-5^*. Taxonomic assignments were determined using the Lowest Common Ancestor (LCA) algorithm implemented in MEGAN (version 6) [24].

Gene abundance estimates were aggregated to generate abundance tables at the kingdom, phylum, class, order, family, genus, and species levels. Taxonomic profiles were summarized to generate relative abundance plots, and differences in microbial community composition and taxonomic abundance between groups were evaluated using multivariate and differential abundance analyses.

### Functional analysis

Functional annotation was performed by aligning predicted protein sequences against the Carbohydrate-Active Enzymes (CAZy) database (version: 2024.03) [25], using DIAMOND (version 0.9.9.110) with the blastp algorithm and an E-value cutoff of *1 × 10^-5^* [23]. For each predicted gene, the best alignment hit was retained for functional annotation. The relative abundance of each carbohydrate-active enzyme was calculated as the sum of the abundances of genes assigned to the corresponding enzyme category.

Differentially abundant carbohydrate-active enzymes between the pre-treatment and post-treatment groups were identified based on their relative abundances and visualized using relative abundance plots.

### Statistical analysis

Continuous clinical variables are presented as mean ± standard deviation (SD). Paired *t*-tests were used to compare BOP and PI between pre-treatment and post-treatment samples. Differences in microbial species abundance and carbohydrate-active enzyme abundance between the two groups were assessed using the Mann–Whitney U test. Differences in microbial community composition were evaluated using analysis of similarities (ANOSIM) implemented in the R package vegan (version 2.3-5). ANOSIM is a nonparametric test based on ranked dissimilarity. All statistical tests were two-sided, and p-values < 0.05 were considered statistically significant.

## 3. RESULTS

### Clinical characteristics of the cohort

The study cohort consisted of 39 patients (21 males and 18 females) with a mean age of 56.36 ± 13.24 years (Table 1). Periodontal therapy was associated with significant improvements in clinical periodontal status, as evidenced by reductions in both BOP (36.53% ± 26.39% vs. 29.19% ± 23.95%, p = 0.013) and PI (65.58% ± 26.93% vs. 47.27% ± 26.89%, p < 0.001). The number of teeth remained stable during the study period (25.34 ± 3.43), indicating that no teeth were lost following treatment.

**Table 1.**
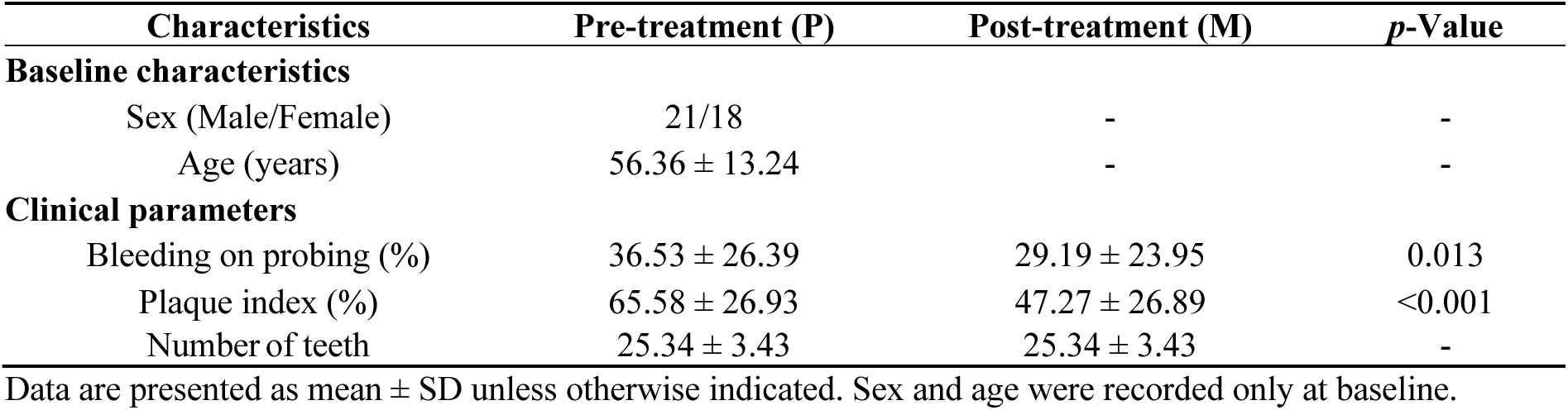
Characteristics of the study cohort of 39 patients.

| Characteristics | Pre-treatment (P) | Post-treatment (M) | $p$-Value |
| --- | --- | --- | --- |
| **Baseline characteristics** |  |  |  |
| Sex (Male/Female) | 21/18 | - | - |
| Age (years) | $56.36 \pm 13.24$ | - | - |
| **Clinical parameters** |  |  |  |
| Bleeding on probing (%) | $36.53 \pm 26.39$ | $29.19 \pm 23.95$ | 0.013 |
| Plaque index (%) | $65.58 \pm 26.93$ | $47.27 \pm 26.89$ | $<0.001$ |
| Number of teeth | $25.34 \pm 3.43$ | $25.34 \pm 3.43$ | - |

Data are presented as mean ± SD unless otherwise indicated. Sex and age were recorded only at baseline.

### Non-redundant gene diversity in pre- and post-treatment plaque samples

A total of 3,177,278 non-redundant genes were identified from the 78 dental plaque samples. As shown in Figure 1A, the median number of non-redundant genes per sample was 837,628 in the pre-treatment (P) group and 894,310 in the post-treatment (M) group. Although the P group exhibited a lower median number of non-redundant genes per sample, it showed greater inter-individual variation, as reflected by a wider interquartile range (IQR: 478,956-998,273) compared with the M group (IQR: 799,680-1,000,594).

**Figure 1.**
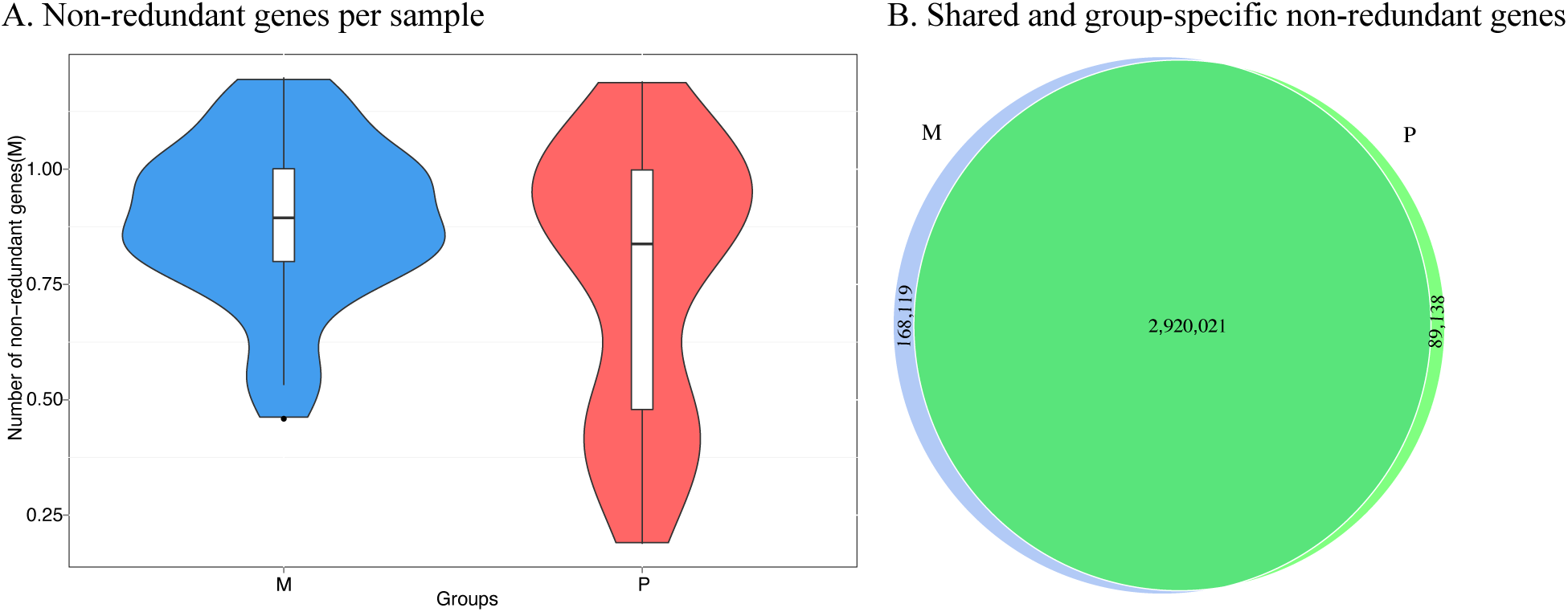
Comparison of non-redundant gene content between pre- and post-treatment dental plaque microbiomes. (A) Distribution of the number of non-redundant genes detected per sample in the post-treatment (M) and pre-treatment (P) groups. Violin plots show the density distribution of gene richness, with embedded boxplots indicating the median and interquartile range. (B) Overlap of non-redundant genes identified in the two groups. Numbers indicate genes unique to each group and those shared between groups.

To assess the overall gene repertoire associated with periodontal treatment, we compared the non-redundant gene catalogues identified in the two groups. A total of 2,920,021 non-redundant genes were shared between the P and M groups, while 89,138 and 168,119 genes were uniquely detected in the pre-treatment and post-treatment groups, respectively (Figure 1B). The large proportion of shared genes indicates that the core functional potential of the dental plaque microbiome was largely conserved following treatment. However, the presence of group-specific genes suggests that periodontal therapy was associated with shifts in microbial composition and functional capacity that may contribute to treatment-induced remodeling of the oral microbiome.

### Microbial species richness and community composition

A total of 12,353 microbial species were identified across the 78 dental plaque samples through taxonomic annotation. The post-treatment (M) group exhibited a higher microbial richness than the pre-treatment (P) group, with an average of 4,343 and 3,581 species detected per sample, respectively (Figure 2A). The median number of species per sample was 4,308 in the M group and 3,586 in the P group.

**Figure 2.**
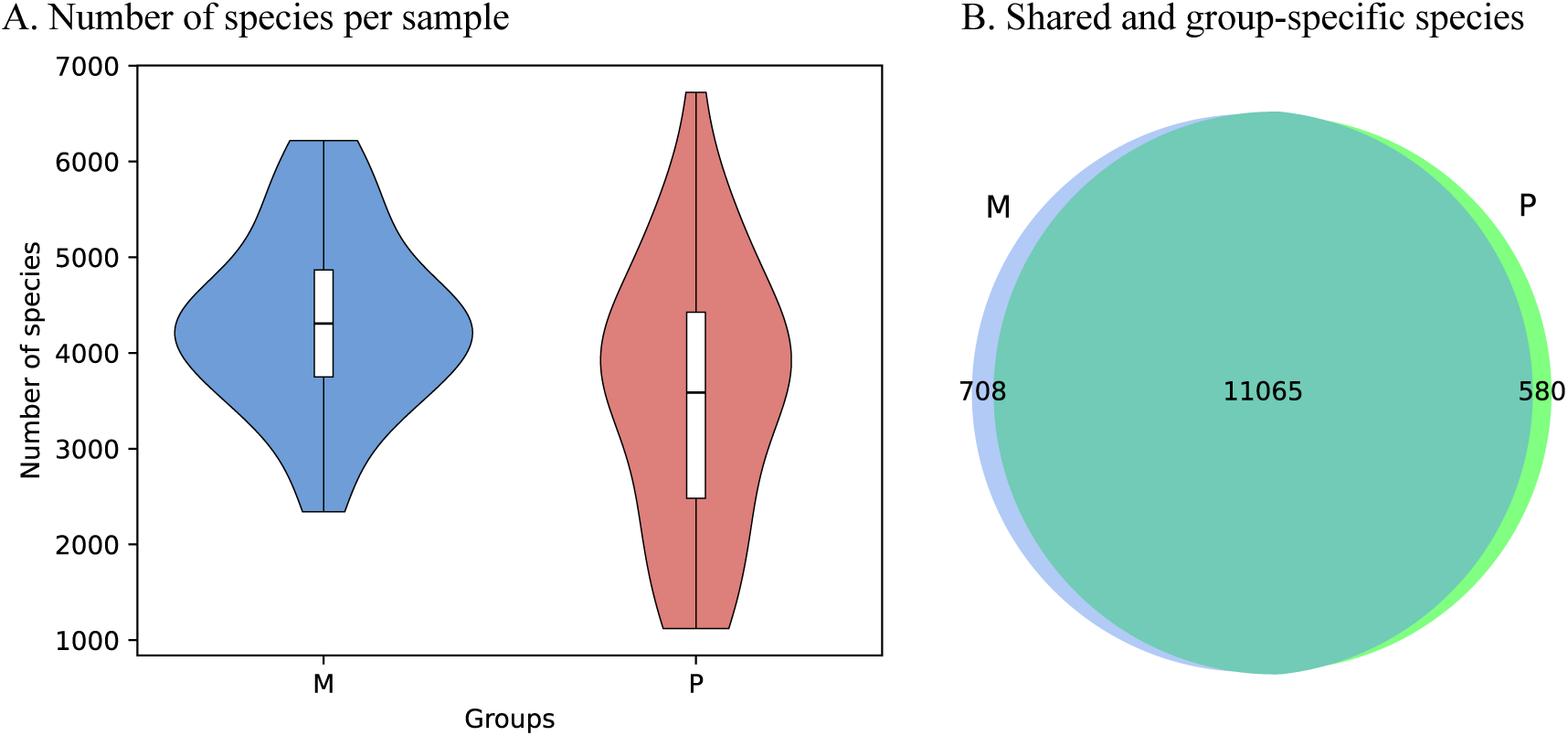
Comparison of species between pre- and post-treatment dental plaque microbiomes. (A) Distribution of the number of species per sample detected in the post-treatment (M) and pre-treatment (P) groups. Violin plots show the density distribution of species richness, with embedded boxplots indicating the median. (B) Overlap of species identified in the two groups. Numbers indicate species unique to each group and those shared between groups.

Greater inter-individual variation was similarly observed in the P group, as indicated by its wider interquartile range (2,481-4,425) compared with the M group (3,750-4,868).

Comparison of species repertoires between groups revealed that most species were shared between pre- and post-treatment samples, while a subset was uniquely detected in each group (Figure 2B). Post-treatment samples exhibited greater microbial richness, accompanied by changes in community composition.

ANOSIM was performed to assess differences in species-level microbial community composition between the M and P groups (Figure 3). The analysis identified a statistically significant shift in community structure following treatment (ANOSIM R = 0.138, P = 0.001). The relatively low R statistic indicates that the magnitude of this shift was modest and that considerable overlap remained between groups. Nevertheless, the significant p-value supports the presence of treatment-associated changes in species composition, consistent with the increased microbial richness and altered species repertoire observed in post-treatment samples.

**Figure 3.**
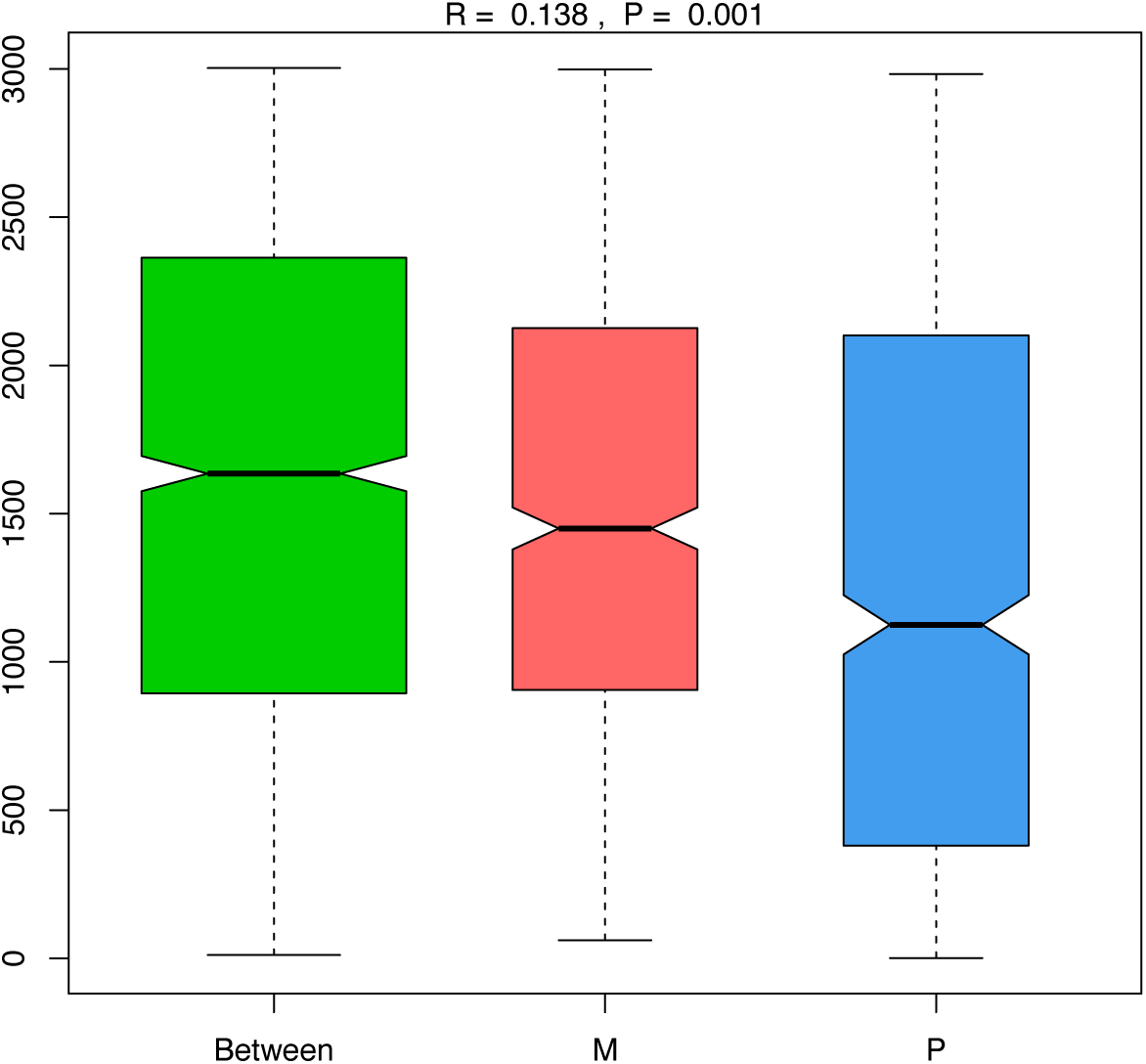
Analysis of Similarities (ANOSIM) of species-level microbial community composition in pre-treatment (P) and post-treatment (M) samples. Boxplots show rank dissimilarities for between-group (Between) and within-group comparisons. The results (R = 0.138, p-value = 0.001) indicate a significant difference in species composition between groups.

### Treatment-associated changes in bacterial species

Figure 4 presents the relative abundance of the ten most abundant bacterial species in the pre- and post-treatment groups. Overall, periodontal therapy was associated with a shift in the relative composition of the dental plaque microbiome, with the post-treatment microbiome showing a greater contribution from *Actinomyces* species, whereas the relative abundance of several disease-associated taxa, including *Escherichia coli*, decreased. Recent studies have identified *E. coli* as a potential contributor to periodontal disease [26–28], making its marked reduction after treatment particularly noteworthy. Although these dominant taxa accounted for the largest treatment-associated differences, they collectively represented less than 9% of the total microbial abundance in either group. More than 91% of the microbiome consisted of lower-abundance species. This finding is consistent with the large numbers of shared non-redundant genes and microbial species identified in the preceding analyses.

**Figure 4.**
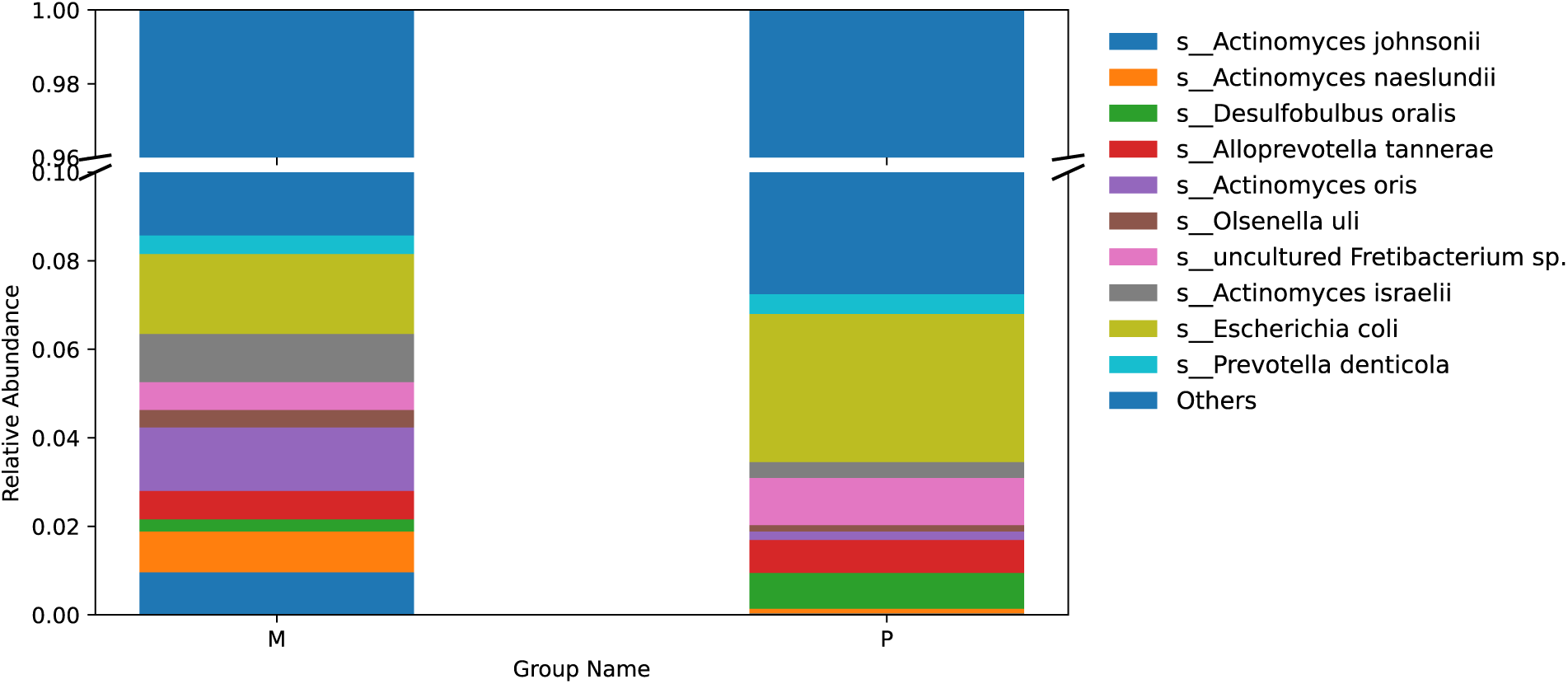
Relative abundance of the ten most abundant bacterial species in pre-treatment (P) and post-treatment (M) dental plaque microbiomes. Relative abundances were averaged across samples within each group and are presented as stacked bar plots. Species names are shown on the right, and the proportion of each species within the total microbial community is indicated on the y-axis.

To further characterize treatment-associated taxonomic changes, the median abundance of selected bacterial species in the pre- and post-treatment groups, together with fold changes and statistical comparisons, is presented in Appendix Table A1. Several established and emerging periodontal pathogens exhibited reduced abundance following treatment. Notably, the keystone pathogen *Porphyromonas gingivalis* and the Red Complex member *Tannerella forsythia* were significantly reduced in post-treatment samples, consistent with previous studies demonstrating suppression of major periodontal pathogens following SRP. In addition, *E. coli* and *Burkholderia multivorans*, opportunistic species associated with dysbiotic microbial communities, were also significantly reduced following therapy.

Conversely, several species commonly associated with oral health and early biofilm formation increased after treatment, including *Streptococcus cristatus*, *Actinomyces naeslundii*, *Actinomyces oris*, *Actinomyces johnsonii*, *Actinomyces israelii*, and *Actinomyces gerencseriae* [29, 30]. Of particular interest, *S. cristatus* has been shown to inhibit *P. gingivalis* fimbrial gene expression and colonization [11, 31]. In contrast, *Filifactor alocis*, an emerging periodontal pathogen, was not reduced following treatment. The persistence of this organism despite clinical improvement suggests that certain disease-associated taxa may remain after successful periodontal therapy, consistent with recent reports that oral microbiome dysbiosis can persist despite treatment-induced remission of periodontal disease.

### Treatment-associated functional remodeling of the oral microbiome

To investigate whether the functional potential of the dental plaque microbiome changed following non-surgical periodontal therapy, predicted microbial genes were annotated against the Carbohydrate-Active Enzymes (CAZy) database. Comparison of enzyme profiles revealed significant differences in several carbohydrate-active enzymes between pre- and post-treatment samples (Figure 5 and Appendix Table A2). In particular, multiple glycosyltransferases, including hyaluronan synthase (EC 2.4.1.212) and β-1,4-mannan synthase, as well as several additional carbohydrate biosynthetic enzymes, exhibited significantly greater abundance in post-treatment samples. Conversely, Dol-P-Man α-1,4-mannosyltransferase showed reduced abundance following treatment.

**Figure 5.**
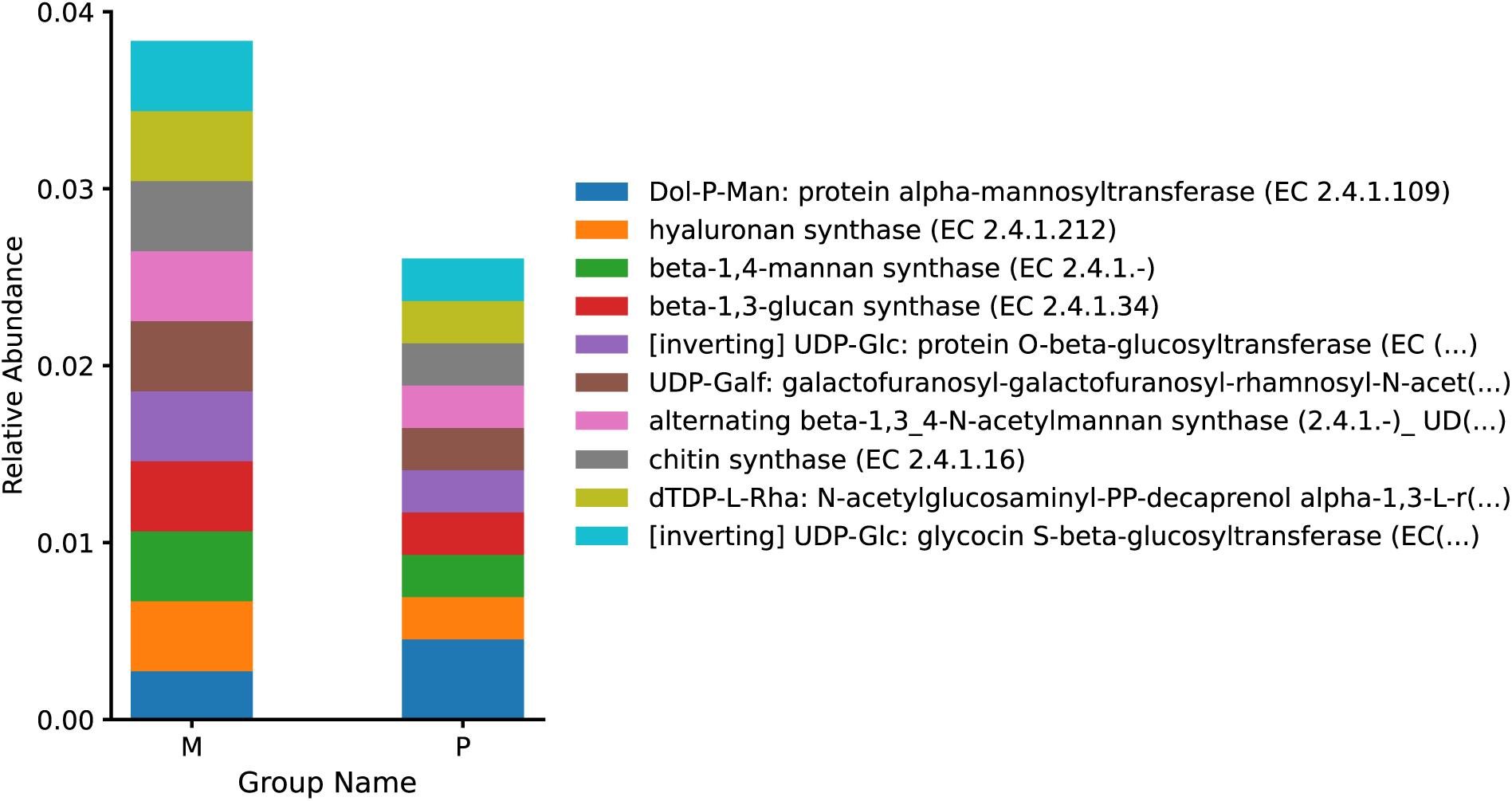
Relative abundance of carbohydrate-active enzymes (CAZy) identified in pre- and post-treatment dental plaque microbiomes. Predicted microbial genes were annotated against the Carbohydrate-Active Enzymes (CAZy) database. Stacked bar plots compare the relative abundance of the ten most abundant CAZy-associated enzyme activities (EC numbers) between the pre-treatment (P) and post-treatment (M) groups. Post-treatment samples exhibited increased abundance of multiple glycosyltransferases and other carbohydrate-active enzymes.

The increased abundance of carbohydrate-active enzymes after treatment paralleled the enrichment of *Actinomyces* and other health-associated commensal bacteria observed in the earlier taxonomic analyses. Together, these findings indicate treatment-associated changes in both microbial composition and predicted functional potential.

## 4. DISCUSSION

Successful periodontal therapy aims not only to suppress periodontal pathogens but also to restore a balanced and functionally resilient oral microbial ecosystem. Using whole-metagenome shotgun sequencing, we comprehensively characterized taxonomic and predicted functional changes in the dental plaque microbiome following SRP. Compared with previous studies based on checkerboard DNA–DNA hybridization, qPCR, or 16S rRNA sequencing, shotgun metagenomics provides species-level resolution while simultaneously enabling functional characterization of the oral microbiome. Our findings demonstrate that successful SRP is associated with significant clinical improvement accompanied by increased microbial richness, shifts in community composition, and coordinated taxonomic and predicted functional remodeling of the dental plaque microbiome.

One notable observation was the increase in microbial richness following treatment. Although the majority of genes and microbial species were shared between pre- and post-treatment samples, the post-treatment microbiome contained a greater number of non-redundant genes and microbial species per sample. Together with the significant ANOSIM results, these findings suggest that periodontal therapy reshapes the microbial community while preserving much of its core repertoire. This observation is consistent with recent longitudinal studies demonstrating that periodontal treatment shifts the oral microbiome toward a healthier ecological state rather than completely replacing the existing microbial community.

At the species level, periodontal therapy reduced several well-established periodontal pathogens, including *P. gingivalis* and *T. forsythia*, consistent with numerous previous reports. In addition, opportunistic organisms such as *E. coli* and *B. multivorans* were also significantly reduced following treatment. Although *E. coli* has traditionally been regarded as a transient member of the oral microbiota, increasing evidence suggests that it may contribute to periodontal disease. Experimental studies have shown that lipopolysaccharides (LPS) derived from *P. gingivalis* and *E. coli* stimulate inflammatory responses through distinct Toll-like receptor pathways, with *P. gingivalis* LPS signaling primarily through TLR2 and *E. coli* LPS through TLR4 [26]. Both LPS preparations also induce the expression of inflammatory mediators, including TNF-α and VCAM-1, although *E. coli* LPS has been reported to elicit a stronger inflammatory response [27]. Together with recent clinical evidence demonstrating increased abundance of *E. coli* in periodontitis and reductions following periodontal therapy [28], our findings provide additional metagenomic evidence supporting its association with periodontal dysbiosis.

Interestingly, although *E. coli* abundance decreased significantly following therapy, it was detected in all metagenomic samples before and after treatment. This observation differs from previous PCR-based studies reporting lower detection frequencies [32] and may reflect the greater sensitivity and broader taxonomic coverage of whole-metagenome shotgun sequencing compared with PCR-based detection methods.

Conversely, several health-associated species increased following treatment, particularly multiple *Actinomyces* species and *S. cristatus*. These organisms are well-recognized early colonizers that contribute to the establishment of healthy multispecies dental biofilms. Previous studies have shown that *S. cristatus* suppresses *P. gingivalis* colonization by inhibiting expression of its fimbrial genes, while *Actinomyces* species contribute to biofilm maturation through interactions with other commensal organisms. The enrichment of these taxa following SRP therefore suggests ecological recovery of the dental plaque microbiome rather than simply elimination of periodontal pathogens. Interestingly, *F. alocis* was not significantly reduced after treatment, indicating that certain disease-associated organisms may persist despite clinical remission.

An important contribution of this study is the integration of functional analysis with species-level taxonomic profiling. Functional annotation using the CAZy database demonstrated treatment-associated differences in several carbohydrate-active enzymes, particularly glycosyltransferases involved in carbohydrate metabolism and extracellular polysaccharide biosynthesis. These functional changes paralleled the enrichment of *Actinomyces* species and *S. cristatus*, suggesting that recovery of health-associated microbial communities is accompanied by remodeling of the predicted metabolic capacity of the dental plaque microbiome. Rather than reflecting only suppression of pathogenic organisms, successful periodontal therapy appears to promote restoration of microbial functions associated with biofilm development and ecological stability.

This study has several strengths. The paired pre- and post-treatment design minimized inter-individual variability, while shotgun metagenomic sequencing provided substantially greater taxonomic resolution than 16S rRNA sequencing and enabled simultaneous characterization of microbial genes, species composition, and predicted functional potential. Nevertheless, several limitations should be acknowledged. The cohort size was modest and samples were collected at a single post-treatment time point, limiting assessment of long-term microbiome dynamics. Functional analyses were based on genomic annotation rather than direct measurements of gene expression or metabolic activity. Future studies incorporating larger longitudinal cohorts together with metatranscriptomic or metabolomic analyses will further clarify the functional mechanisms underlying microbiome recovery following periodontal therapy.

## 5. CONCLUSIONS

This study demonstrates that non-surgical periodontal therapy (SRP) is associated with coordinated taxonomic and predicted functional remodeling of the dental plaque microbiome. Whole-metagenome shotgun sequencing revealed increased microbial richness and significant shifts in community composition following treatment, characterized by suppression of disease-associated taxa and enrichment of health-associated early colonizers. Functional annotation further identified treatment-associated changes in carbohydrate-active enzymes, suggesting that microbial recovery involves not only taxonomic restructuring but also alterations in the predicted metabolic capacity of the oral microbiome. Collectively, these findings provide a more comprehensive understanding of microbiome remodeling following periodontal therapy and demonstrate the value of whole-metagenome shotgun sequencing for investigating both microbial composition and functional responses to treatment.

## Author Contributions

H.X. contributed to the conception and design of the study. Q.W. and H.X. performed data analysis and prepared Tables and Figures. B.-Y.W. prepared Table 1 and collected dental plaque samples. D.W. was involved in data and statistical analyses. All authors have read and agreed to the published version of the manuscript.

## Institutional Review Board Statement

The study was conducted in accordance with the Declaration of Helsinki and approved by the Committee for the Protection of Human Subjects of the University of Texas Health Science Center at Houston (IRB number: HSC-DB-17-0636 and 17 August 2017).

## Informed Consent Statement

Informed consent was obtained from all subjects involved in the study.

## Data Availability Statement

Our metagenomic sequencing data have been deposited in the NCBI Sequence Read Archive (SRA) under accession PRJNA1482867 and will be made public upon paper acceptance.

## Abbreviations

The following abbreviations are used in this manuscript:

CAL: Clinical attachment level
IQR: Interquartile range
PD: Probing depth
SRP: Scaling and root planing

## APPENDIX A

**Appendix Table A1.**
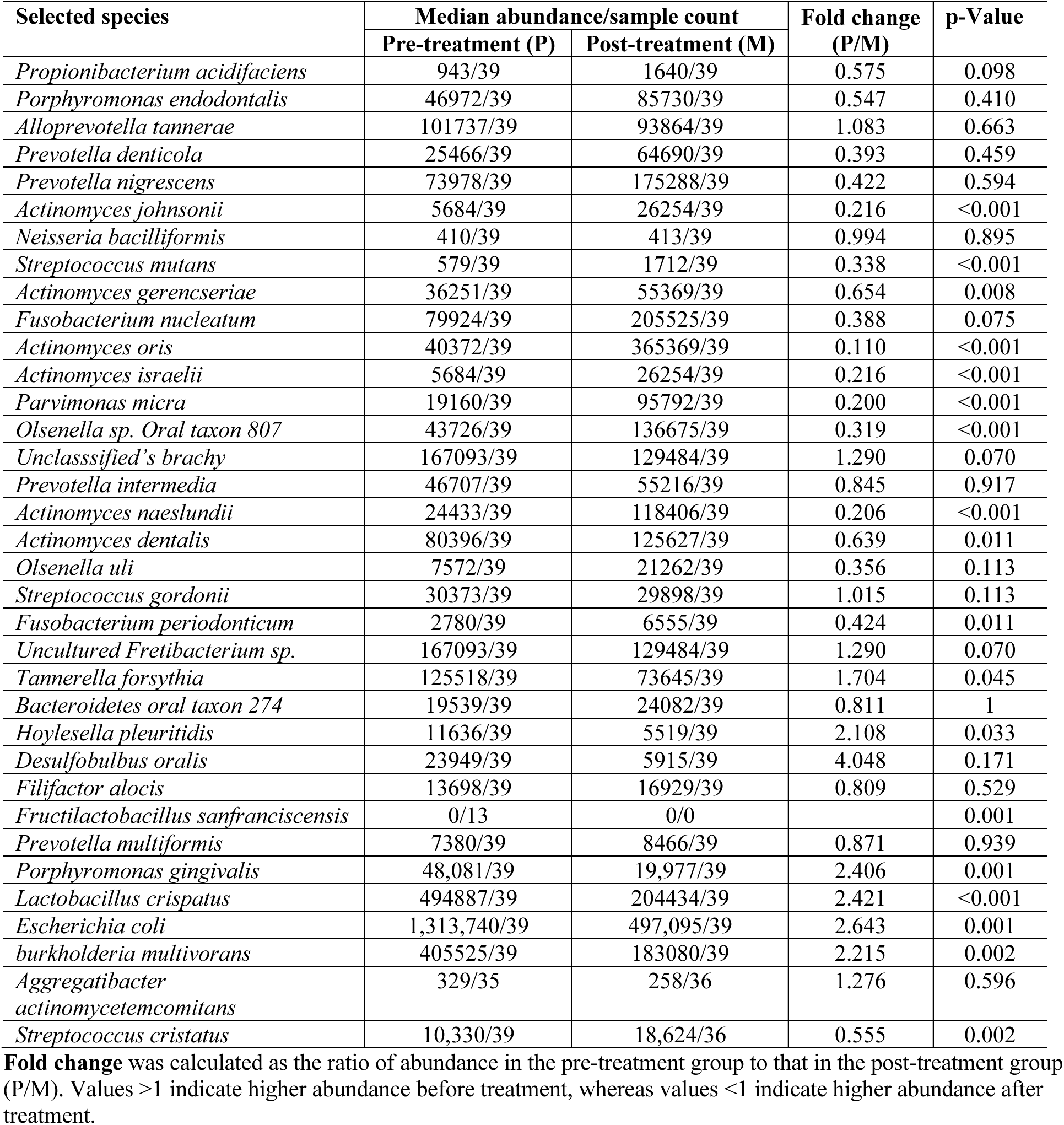
Comparison of median bacterial species abundance between pre- and post-treatment dental plaque microbiomes.

**Appendix Table A2.** Differentially abundant carbohydrate-active enzymes (CAZy) identified in pre- and post-treatment dental plaque microbiomes.

| Enzyme (EC number) | Median abundance (log10+1) |  | Median fold change (P/M) | p-Value |
| --- | --- | --- | --- | --- |
|  | Pre-treatment (P) | Post-treatment (M) |  |  |
| Dol-P-Man alpha-1,4-mannosyltransferase (EC 2.4.1.-) | 1.88 (36) | 1.55 (38) | 1.777 (36) | 0.003 |
| hyaluronan synthase (EC 2.4.1.212) | 5.02 (39) | 5.38 (39) | 0.686 (39) | <0.001 |
| beta-1,4-mannan synthase (EC 2.4.1.-) | 5.02 (39) | 5.38 (35) | 0.686 (39) | <0.001 |
| beta-1,3-glucanase (EC 3.2.1.-) | 0.00 (13) | 1.20 (39) | 0.000 (13) | <0.001 |
| [inverting] UDP-Glc: protein O-beta-glucosyltransferase (EC 2.4.1.-) | 5.02 (39) | 5.38 (39) | 0.686 (39) | <0.001 |
| UDP-Galf: galactofuranosyl-galactofuranosyl-rhamnosyl-N-acetylglucosaminyl-PP-decaprenol beta-1,5_1,6-galactofuranosyltransferase (EC 2.4.1.288) | 5.02 (39) | 5.38 (39) | 0.686 (39) | <0.001 |
| alternating beta-1,3_4-N-acetylmannan synthase (2.4.1.-)_UDP-GlcA: N-acetylglucosaminyl-proteoglycan beta-1,4-glucuronosyltransferase (EC 2.4.1.225) | 5.02 (39) | 5.38 (39) | 0.686 (39) | <0.001 |
| chitin synthase (EC 2.4.1.16) | 5.02 (39) | 5.38 (39) | 0.686 (39) | <0.001 |
| dTDP-L-Rha: N-acetylglucosaminyl-PP-decaprenol alpha-1,3-L-rhamnosyltransferase (EC 2.4.1.289) | 5.02 (39) | 5.38 (39) | 0.686 (39) | <0.001 |
| [inverting] UDP-Glc: glycocin S-beta-glucosyltransferase (EC 2.4.1.-) | 5.02 (39) | 5.38 (39) | 0.686 (39) | <0.001 |

Sample count is shown in parentheses within the cell

## REFERENCES

1. Kassebaum NJ, Bernabe E, Dahiya M, Bhandari B, Murray CJ, Marcenes W. Global burden of severe periodontitis in 1990-2010: a systematic review and meta-regression. Journal of dental research. 2014;93(11):1045–53.

2. Burt B, Research S, Therapy Committee of the American Academy of P. Position paper: epidemiology of periodontal diseases. Journal of periodontology. 2005;76(8):1406–19.

3. Petersen PE, Ogawa H. The global burden of periodontal disease: towards integration with chronic disease prevention and control. Periodontology 2000. 2012;60(1):15–39.

4. Eke PI, Thornton-Evans GO, Wei L, Borgnakke WS, Dye BA, Genco RJ. Periodontitis in US Adults: National Health and Nutrition Examination Survey 2009-2014. Journal of the American Dental Association. 2018;149(7):576–88 e6.

5. Socransky SS, Haffajee AD, Teles R, Wennstrom JL, Lindhe J, Bogren A, et al. Effect of periodontal therapy on the subgingival microbiota over a 2-year monitoring period. I. Overall effect and kinetics of change. J Clin Periodontol. 2013;40(8):771–80.

6. Colombo AP, Bennet S, Cotton SL, Goodson JM, Kent R, Haffajee AD, et al. Impact of periodontal therapy on the subgingival microbiota of severe periodontitis: comparison between good responders and individuals with refractory periodontitis using the human oral microbe identification microarray. Journal of periodontology. 2012;83(10):1279–87.

7. Yama K, Nishimoto Y, Kumagai K, Jo R, Harada M, Maruyama Y, et al. Dysbiosis of oral microbiome persists after dental treatment-induced remission of periodontal disease and dental caries. mSystems. 2023;8(5):e0068323.

8. Johnston W, Rosier BT, Carda-Dieguez M, Paterson M, Watson P, Piela K, et al. Longitudinal changes in subgingival biofilm composition following periodontal treatment. Journal of periodontology. 2023;94(9):1065–77.

9. Simpson A, Johnston W, Carda-Dieguez M, Mira A, Easton C, Henriquez FL, et al. Periodontal treatment causes a longitudinal increase in nitrite-producing bacteria. Mol Oral Microbiol. 2024.

10. Baima G, Ferrocino I, Del Lupo V, Colonna E, Thumbigere-Math V, Caviglia GP, et al. Effect of Periodontitis and Periodontal Therapy on Oral and Gut Microbiota. Journal of dental research. 2024;103(4):359–68.

11. Wang Q, Wang B-Y, Pratap S, Xie H. Oral microbiome associated with differential ratios of *Porphyromonas gingivalis* and *Streptococcus cristatus*. Microbiology Spectrum. 2024;12(2).

12. Wang Q, Wang B-Y, Williams SN, Xie H. Diversity and Characteristics of the Oral Microbiome Associated with Self-Reported Ancestral/Ethnic Groups. International Journal of Molecular Sciences. 2024;25(24):13303.

13. Newman M, Takei, H., Klokkevold, H., & Carranza, F. Newman and Carranza’s Clinical Periodontology. 2018.

14. Papapanou PN, Sanz M, Buduneli N, Dietrich T, Feres M, Fine DH, et al. Periodontitis: Consensus report of workgroup 2 of the 2017 World Workshop on the Classification of Periodontal and Peri-Implant Diseases and Conditions. Journal of periodontology. 2018;89 Suppl 1:S173–S82.

15. Tonetti MS, Greenwell H, Kornman KS. Staging and grading of periodontitis: Framework and proposal of a new classification and case definition. J Clin Periodontol. 2018;45 Suppl 20:S149–S61.

16. Wang BY, Wu J, Lamont RJ, Lin X, Xie H. Negative correlation of distributions of Streptococcus cristatus and Porphyromonas gingivalis in subgingival plaque. Journal of clinical microbiology. 2009;47(12):3902–6.

17. Chen S. fastp 1.0: An ultra-fast all-round tool for FASTQ data quality control and preprocessing. iMeta. 2025;4(5).

18. Langmead B, Salzberg SL. Fast gapped-read alignment with Bowtie 2. Nat Methods. 2012;9(4):357–9.

19. Li D, Luo R, Liu C-M, Leung C-M, Ting H-F, Sadakane K, et al. MEGAHIT v1.0: A fast and scalable metagenome assembler driven by advanced methodologies and community practices. Methods. 2016;102:3–11.

20. Zhu W, Lomsadze A, Borodovsky M. Ab initio gene identification in metagenomic sequences. Nucleic acids research. 2010;38(12):e132.

21. Li W, Godzik A. Cd-hit: a fast program for clustering and comparing large sets of protein or nucleotide sequences. Bioinformatics. 2006;22(13):1658–9.

22. Fu L, Niu B, Zhu Z, Wu S, Li W. CD-HIT: accelerated for clustering the next-generation sequencing data. Bioinformatics. 2012;28(23):3150–2.

23. Buchfink B, Xie C, Huson DH. Fast and sensitive protein alignment using DIAMOND. Nat Methods. 2015;12(1):59–60.

24. Le Chatelier E, Nielsen T, Qin J, Prifti E, Hildebrand F, Falony G, et al. Richness of human gut microbiome correlates with metabolic markers. Nature. 2013;500(7464):541–6.

25. Cantarel BL, Coutinho PM, Rancurel C, Bernard T, Lombard V, Henrissat B. The Carbohydrate-Active EnZymes database (CAZy): an expert resource for Glycogenomics. Nucleic acids research. 2009;37(Database issue):D233–8.

26. Palaska I, Gagari E, Theoharides TC. The effects of P. gingivalis and E. coli LPS on the expression of proinflammatory mediators in human mast cells and their relevance to periodontal disease. J Biol Regul Homeost Agents. 2016;30(3):655–64.

27. Liu R, Desta T, Raptis M, Darveau RP, Graves DT. P. gingivalis and E. coli lipopolysaccharides exhibit different systemic but similar local induction of inflammatory markers. Journal of periodontology. 2008;79(7):1241–7.

28. Veras EL, Castro Dos Santos N, Souza JGS, Figueiredo LC, Retamal-Valdes B, Barao VAR, et al. Newly identified pathogens in periodontitis: evidence from an association and an elimination study. J Oral Microbiol. 2023;15(1):2213111.

29. Kolenbrander PE, Palmer RJ, Periasamy S, Jakubovics NS. Oral multispecies biofilm development and the key role of cell–cell distance. Nature Reviews Microbiology. 2010;8(7):471–80.

30. Mark Welch JL, Rossetti BJ, Rieken CW, Dewhirst FE, Borisy GG. Biogeography of a human oral microbiome at the micron scale. Proceedings of the National Academy of Sciences. 2016;113(6):E791–E800.

31. Xie H, Cook GS, Costerton JW, Bruce G, Rose TM, Lamont RJ. Intergeneric Communication in Dental Plaque Biofilms. Journal of Bacteriology. 2000;182(24):7067–9.

32. Estemalik J, Demko C, Bissada NF, Joshi N, Bodner D, Shankar E, et al. Simultaneous Detection of Oral Pathogens in Subgingival Plaque and Prostatic Fluid of Men With Periodontal and Prostatic Diseases. Journal of periodontology. 2017;88(9):823–9.

